# Research on the mental health status of frontline medical staff during the normalization of the COVID-19 pandemic

**DOI:** 10.1101/2023.01.09.23284361

**Authors:** Ning Sun, Laiyou Li, Jinmei Xu, Shuping zhou, Chaoyan Fan, Hongyu Li, Shuang Yang

**Affiliations:** Ningbo College of Health Sciences, Ningbo, P.R. China; The affiliated people’s hospital of Ningbo University, Ningbo, P.R. China

**Author notes:** Jinmei Xu and Shuping Zhou were co-corresponding author, Corresponding authors at: Jinmei Xu: Ningbo College of Health Sciences, Ningbo, P. R. China.& Shuping Zhou: Ningbo College of Health Sciences, Ningbo, P. R. China. 8615888174602. 8615888121394. 8613957880925. 8618858234686. Ning Sun and Laiyou Li were the co-first author.

**Keywords:** Medical staff, New coronavirus pneumonia, SCL-90 scale, Psychological health, Influencing factors

## Abstract

**Objective:** This study aimed to investigate the relationship between personality characteristics and psychological health of hospitals’ frontline medical staff and provide a basis and reference for targeted psychological health education for frontline medical staff and for the staff of related departments to formulate relevant policies.

**Methods:** The self-evaluation scale of symptoms (SCL-90) was used to investigate the mental health status of 150 first-line medical staff in Zhejiang Province in response to the new severe acute respiratory syndrome coronavirus 2 (SARS-CoV-2) pneumonia.

**Results:** The average scores of SCL-90 and somatization, obsessive-compulsive, depression, anxiety, hostility, terror, and psychotic factors were significantly higher than those of the normal sample in the first-aid medical staff of Aihu Hubei. The degree of influence on the mental health status of the frontline medical staff in service in Hubei is as follows, from high to low: the degree of suspicion that they may have been infected when new coronavirus pneumonia-related symptoms occur, the degree of fear of being infected and thus bring the infection to their families, and whether they have received a medical check-up recently, as well as a high level of education (both *P*<0.05).

**Conclusion:** The psychological health level of the frontline medical staff is lower than the national norm. In the context of the increasing number of confirmed cases and the new type of coronavirus pneumonia in the absence of any specific curative treatments, the frontline medical staff is under great psychological pressure. It is necessary to institute targeted mental health promotion to relieve the psychological pressure endured by the frontline medical staff, promote their physical and mental health, and better respond to the pandemic in China.

## Introduction

The novel coronavirus pneumonia (NCP) has the new severe acute respiratory syndrome coronavirus 2 (SARS-CoV-2) as its pathogen. On January 12, 2020, the World Health Organization (WHO) officially named this new disease the 2019 coronavirus disease (COVID-19). The first case of COVID-19 occurred in Wuhan on December 12, 2019 [1,2]. Although SARS-CoV-2 shares some similarities with the coronaviruses SARS-CoV and Middle East respiratory syndrome–related coronavirus (MERS-CoV), the rapidly increasing number of cases and the evidence of more human-to-human transmission have shown that SARS-CoV-2 is more contagious than the other two and is a new coronavirus strain that had never been found in human beings [3,4]. The common signs of NCP include fever, cough, shortness of breath, and breathing difficulties. In more severe cases, pneumonia, severe acute respiratory failure, renal failure, and even death can occur [5]. Moreover, there is currently no specific treatment for NCP.

The COVID-19 outbreak can cause not only physical damage to infected people but also serious mental illness. During the COVID-19 outbreak, people’s anxiety and depression levels significantly increased, and their sleep quality deteriorated [6,7]. Notably, medical staff were also prone to such psychological problems [8] due to their significantly increased workload and raised risk of infection. In addition, facing patients’ negative emotions and the fear of transmitting the virus to their family members contributed to the psychological load. A survey of the psychological status of 1,995 Finnish medical staff showed that during the COVID-19 outbreak, 40% of the respondents showed mild to moderate anxiety, and 5% of the medical staff showed severe anxiety [9]. Another report on the mental health impact of COVID-19 on doctors in the United States showed that 67.6% of 1,356 doctors reported a moderate or high degree of emotional distress [10]. The COVID-19 pandemic has been effectively contained [11] in China, owing to the government’s timely action, the cooperation of the masses, and the high-intensity operativeness of the medical staff [12,13]. However, even under the strict prevention and control of COVID-19 policy [14,15], new cases are still arising, and there are asymptomatic infected people in China, making the COVID-19 pandemic more “normal” as time goes by.

On December 7, 2022, the State Council zone spreading mechanism group announced further optimization of medical processes to improve the current medical service by optimizing the medical resources available. They were to ensure that maximizing order and safety in the medical service would not affect routine diagnosis and treatment and the acute management of positive patients. In autumn and winter, the number of patients with respiratory diseases increases. With the added risk introduced by the novel coronavirus infection, the frontline medical staff would be burdened by the task of managing it, with inevitable great psychological pressure. According to data statistics, as of December 12, 2022, there were more than 9.3 million confirmed cases in China, including over 14,000 new cases in a single day. Experts explained that with the first wave of the pandemic after the full release, the peak infection rate would reach 60 percent. In this case, the frontline medical staff would be exposed to a large number of novel coronavirus infections, causing them to worry about being infected due to occupational exposure, and worrying that if they do not work well, they may affect the whole team, thus generating great psychological pressure. In a short time, the number of cases has greatly increased; medical staff must work tirelessly with the consistent pressure of the need to provide treatment, and they may not be able to ensure adequate rest and sleep for themselves. Due to the closed-loop management requirements, some medical staff workers may be separated from their families for a long time, which may also increase their concerns about their family’s health and wellness. Long shifts, intensive work, and constant readiness to deal with large-scale emergencies make the medical staff prone to psychological problems. These, in turn, are associated with many negative outcomes, such as reduced efficiency, loss of productivity, disability, and absenteeism [9]. Given these adverse effects, research is warranted to reveal the potential factors and mechanisms by which mental health of healthcare staff may be improved, and their productivity maintained during the new COVID-19 era.

Previous studies have focused on the changes in the psychological status of medical staff in the early stage of the COVID-19 pandemic and in high-risk areas, while only a few studies have reported the psychological status of the medical staff in low-risk areas and following the “normalization” of the COVID-19 pandemic. Therefore, this study analyzes the status and the influencing factors of psychological health of front-line medical staff in dealing with the NCP to provide an objective basis for prevention and intervention measures. This study aimed to shed light on the relationship between the sociodemographic characteristics of front-line medical workers and their psychological health to provide the basis and reference for targeted mental health education and to formulate appropriate policies for the management of the relevant hospital departments.

## Methods

### Study design

This quantitative study was conducted using a questionnaire to gather psychometric measures.

### Setting and sample

This study used convenient sampling to collect information from 150 front-line health professionals from hospitals operating at different levels in Zhejiang Province between December 8 and 22, 2022. Chinese hospitals are classified into three main levels according to their number of beds. The number of beds in first-level hospitals is below 100, in second-level hospitals it is 101 to 499, and in tertiary hospitals it is above 500. The inclusion criteria for the participants were as follows: 1) front-line medical workers 2) who gave their written informed consent to participate in the study.

### Sample size calculation

The study mainly discusses the correlation between sociodemographic characteristics and psychological stress. Multiple regression analysis was applied. It was estimated that 16 variables may be entered into the model because the sample size was estimated to be at least 10–15 times the variable entered into the model; thus, 160 participants were required. The loss to follow-up rate was calculated at 10%, therefore, the sample size was set to be 176 people.

### Data collection

Two investigators underwent unified training. The data collection was conducted with online interviews. All the front-line health workers worked at hospitals. The investigators explained the research objectives and methods and obtained consent and cooperation from those who met the inclusion criteria. Informed consent was obtained via online interviews, and the research data were collected via an online questionnaire survey. The front-line medics who consented to participate received a link to access the questionnaires. Participants completed the questionnaires immediately upon receipt. To ensure anonymity, nobody except the researchers could see the IP address and any private information about the participants.

### Study measures

Two questionnaires were used in this study. The questionnaire for the sociodemographic characteristics of the medical staff included 16 items: hospital, department, occupation, sex, age, highest level of education, years of service, technical titles, marital status, having children or not, residence, having received training relevant to public health emergencies response or not, having family support while working on the front-line during the pandemic or not, level of fear about getting infected (them and/or their families), level of worry about getting infected when presenting symptoms associated with NCP, and the recent completion of a comprehensive medical check-up. The Symptoms Self-rating Scale (Self-reporting Inventory, SCL-90) includes 10 factors and 90 items, each of which reflects the symptom pain of the medical staff. The characteristics of the symptom distribution can be understood through the factors. The 10 factors include somatization, compulsive symptoms, interpersonal sensitivity, depression, anxiety, hostility, terror, and paranoid, psychotic, and sleep/eating symptoms. Each item adopted the Likert 5-grade score (none, mild, moderate, heavy, and severe). The total score is the sum of the 90 items scored. In previous studies, the homogeneity reliability of the SCL-90 total table was 0.97, the homogeneity reliability of each subscale was also greater than 0.70, the test-retest reliability was greater than 0.7, and the content validity was above 0.80, with good reliability and validity [16].

### Ethical considerations

This study adopted an online questionnaire method, and the researchers conducted the survey after unified training. The research was approved by the Ningbo College of health sciences ethics review board. We approached the medical staff working on the front line and explained to them the research objectives, methods, and other relevant information to facilitate cooperation. After obtaining their agreement, we requested them to sign an online letter of consent, informing them that participation was fully voluntary and that they could choose to withdraw from the research at any time. We then conducted the online survey by sending the questionnaire link to a total of 180 medical staff.

### Statistical methods

SPSS 26 was used to perform data analysis after the logical testing. P <0.05 was considered statistically significant. Mean, standard deviation, and frequency values were used to describe the demographic data of the front-line medical staff; mean and standard deviation values were used to describe the psychological health scores of front-line medical workers.Multiple regression analysis was used to analyze the influence of the medical workers’ socio-demographic data on their levels of psychological health. The total scores of psychological health were chosen as the dependent variables. The 16 items measuring socio-demographic characteristics were the independent variables. The stepwise method was used when the independent variables were entered into the regression analysis. The assignment method of the independent variables was also shown.

## Results

### General information on front-line medical workers

A total of 180 questionnaires were collected online, of which 150 were considered valid, with an effective recovery rate of 83.33%. The detailed demographic information collected is listed in Table 1.

**Table 1.** Demographic data of clinical medical personnel (n=150).

| **project** | **Example number** | **constituent ratio (%)** |
| --- | --- | --- |
| **Hospital level** |  |  |
| Grade 3 A | 125 | 83.3 |
| Grade III B et al | 8 | 5.3 |
| Secondary-level | 14 | 9.3 |
| Grade II B et al | 3 | 2.0 |
| **The Department** |  |  |
| Internal medicine | 36 | 24.0 |
| Surgery | 35 | 23.3 |
| Gynaecology and obstetrics | 4 | 2.7 |
| Paediatrics | 2 | 1.3 |
| ED | 28 | 18.7 |
| Critical care medicine | 25 | 16.7 |
| section for Outpatients | 5 | 3.3 |
| Operating room | 2 | 1.3 |
| Else | 13 | 8.7 |
| **Occupation** |  |  |
| Doctor | 43 | 28.7 |
| Nurse | 107 | 71.3 |
| **Sex** |  |  |
| Man | 56 | 37.3 |
| Woman | 94 | 62.7 |
| **Age distribution (years)** |  |  |
| Under 25 | 4 | 2.7 |
| 26-35 Years old | 84 | 56.0 |
| 36-45 Years old | 41 | 27.3 |
| Over 45 years old | 21 | 14.0 |
| **Highest education** |  |  |
| Special school | 2 | 1.3 |
| Junior college | 10 | 6.7 |
| Undergraduate course | 99 | 66.0 |
| Master's degree or above | 39 | 26.0 |
| **Working life** |  |  |
| 5 Years and below | 20 | 13.3 |
| 6-10 Years | 51 | 34.0 |
| 11-15 Years | 32 | 21.3 |
| 16-20 Years | 21 | 14.0 |
| More than 20 years | 26 | 17.3 |
| **Technical title** |  |  |
| Elementary | 63 | 42.0 |
| Middle rank | 52 | 34.7 |
| Senior | 35 | 23.3 |
| **Marital status** |  |  |
| Unmarried | 33 | 22.0 |
| Married | 111 | 74.0 |
| Dissociaton | 6 | 4.0 |
| **Have children** |  |  |
| have | 109 | 72.7 |
| not have | 41 | 27.3 |
| **Current living situation** |  |  |
| Live alone | 35 | 23.3 |
| With the family | 112 | 74.7 |
| Joint rent | 3 | 2.0 |
| **Have participated in the training on responding to public health emergencies** |  |  |
| Yes | 112 | 74.7 |
| deny | 38 | 25.3 |
| **Whether your family members support you to work on the clinical frontline** |  |  |
| yes | 142 | 94.7 |
| deny | 8 | 5.3 |
| **Worried about the extent to which you and your family will be infected** |  |  |
| Severe | 9 | 6.0 |
| Moderate | 60 | 40.0 |
| Mild | 40 | 26.7 |
| not have | 41 | 27.3 |
| **COVID-19 related symptoms, suspected the degree of infection** |  |  |
| Severe | 67 | 44.7 |
| Moderate | 45 | 30.0 |
| Mild | 22 | 14.7 |
| not have | 16 | 10.7 |
| **Have received recent medical observation** |  |  |
| have | 23 | 15.3 |
| not have | 127 | 84.7 |

### SCL-90 was compared with the national norm

The average score of SCL-90 overall and all the somatic, obsessive-compulsive disorder, depression, anxiety, antagonistic, horror, and psychiatric factors were significantly higher than the norm (P <0.05) [17] (see Table 2).

**Table 2.**
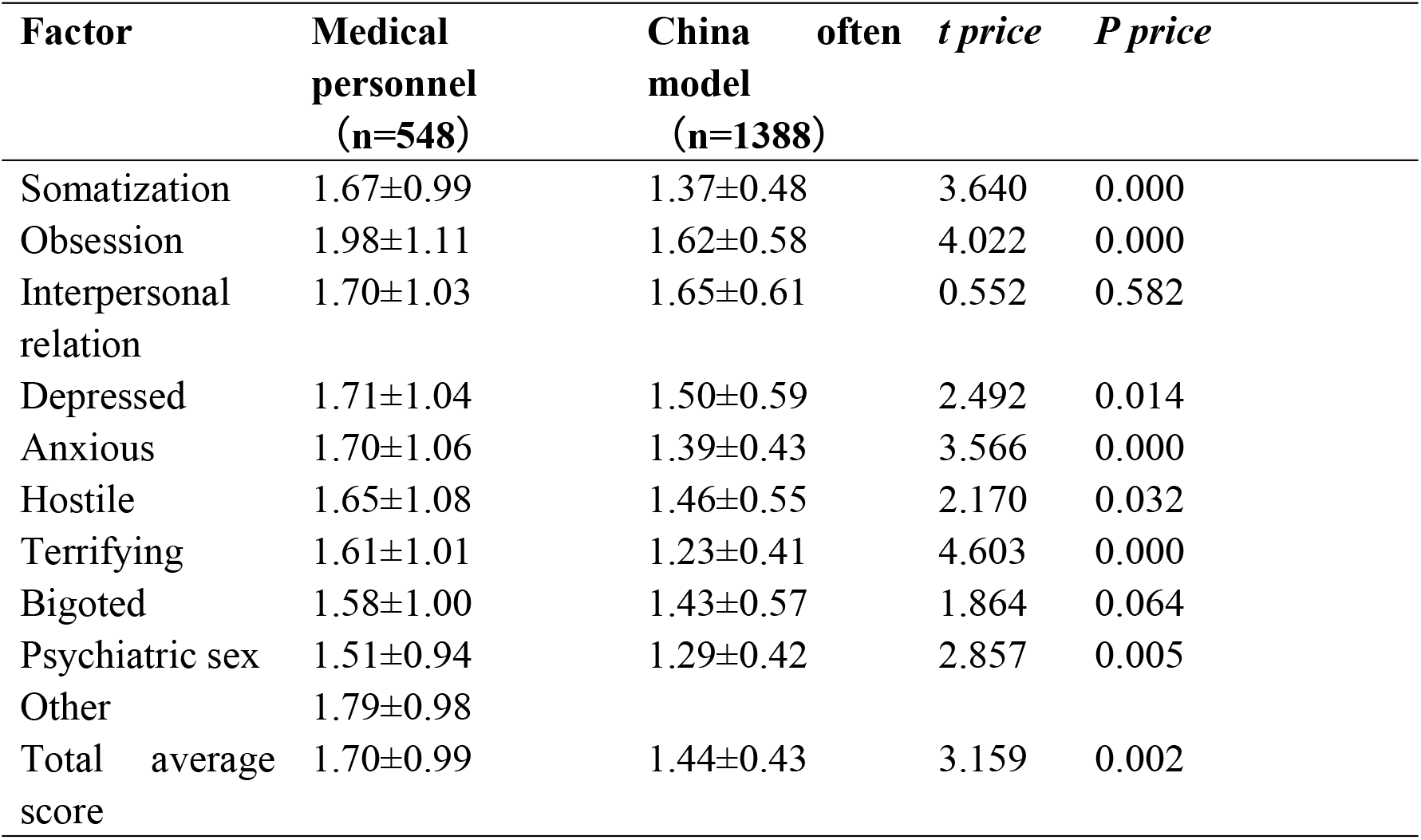
Comparison of F scores (X ± S).

| **Factor** | **Medical**<br>**personnel**<br>**(n=548)** | **China**<br>**often**<br>**model**<br>**(n=1388)** | ***t price*** | ***P price*** |
| --- | --- | --- | --- | --- |
| Somatization | 1.67±0.99 | 1.37±0.48 | 3.640 | 0.000 |
| Obsession | 1.98±1.11 | 1.62±0.58 | 4.022 | 0.000 |
| Interpersonal<br>relation | 1.70±1.03 | 1.65±0.61 | 0.552 | 0.582 |
| Depressed | 1.71±1.04 | 1.50±0.59 | 2.492 | 0.014 |
| Anxious | 1.70±1.06 | 1.39±0.43 | 3.566 | 0.000 |
| Hostile | 1.65±1.08 | 1.46±0.55 | 2.170 | 0.032 |
| Terrifying | 1.61±1.01 | 1.23±0.41 | 4.603 | 0.000 |
| Bigoted | 1.58±1.00 | 1.43±0.57 | 1.864 | 0.064 |
| Psychiatric sex | 1.51±0.94 | 1.29±0.42 | 2.857 | 0.005 |
| Other | 1.79±0.98 |  |  |  |
| Total average<br>score | 1.70±0.99 | 1.44±0.43 | 3.159 | 0.002 |

### Analysis of the factors influencing the psychological status of the clinical medical staff

Multiple linear regression analysis was performed on the total mental health status score and the general data as independent variables; the details of the independent variable assignment methods are shown in Table 2. The results showed that the influencing factors affecting mental health status were, from high to low: the degree of infection when presenting COVID-19-related symptoms, the degree of concern for oneself and one’s family members, the recent medical check-up, and the high level of education (*P* <0.05). The specific results are shown in Table 3.

**Table 3.**
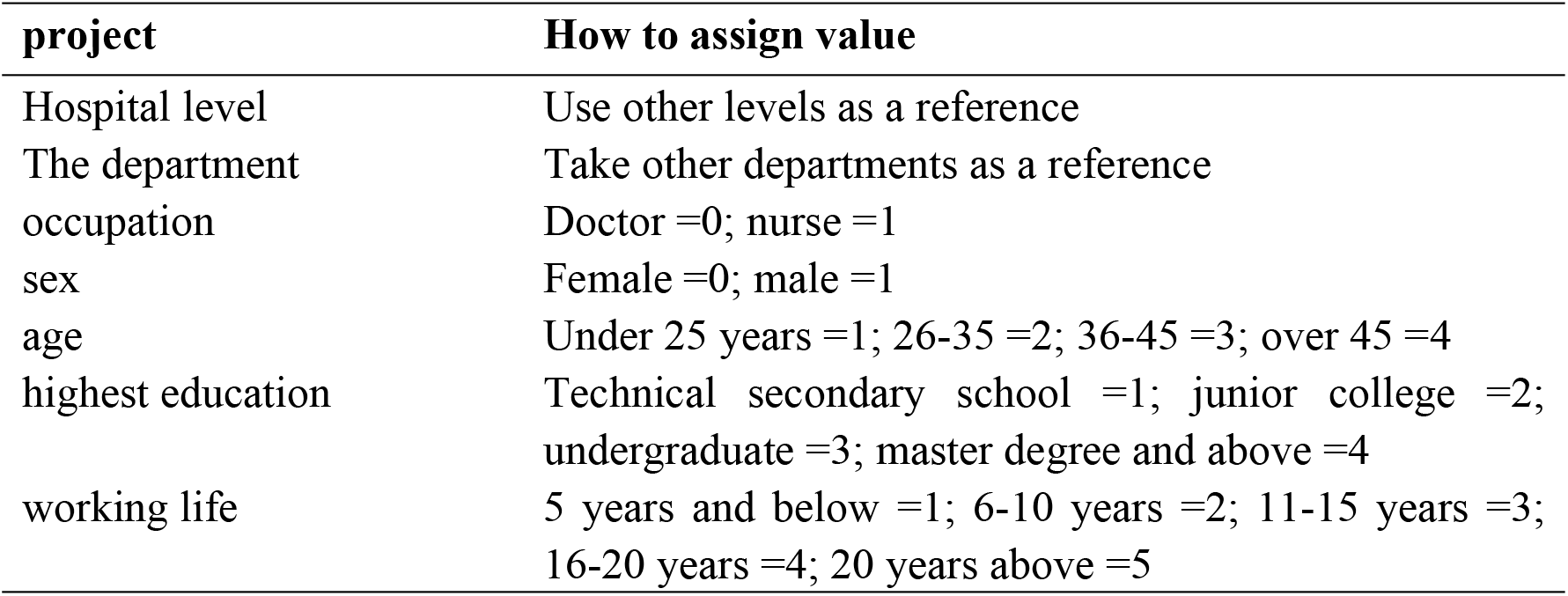
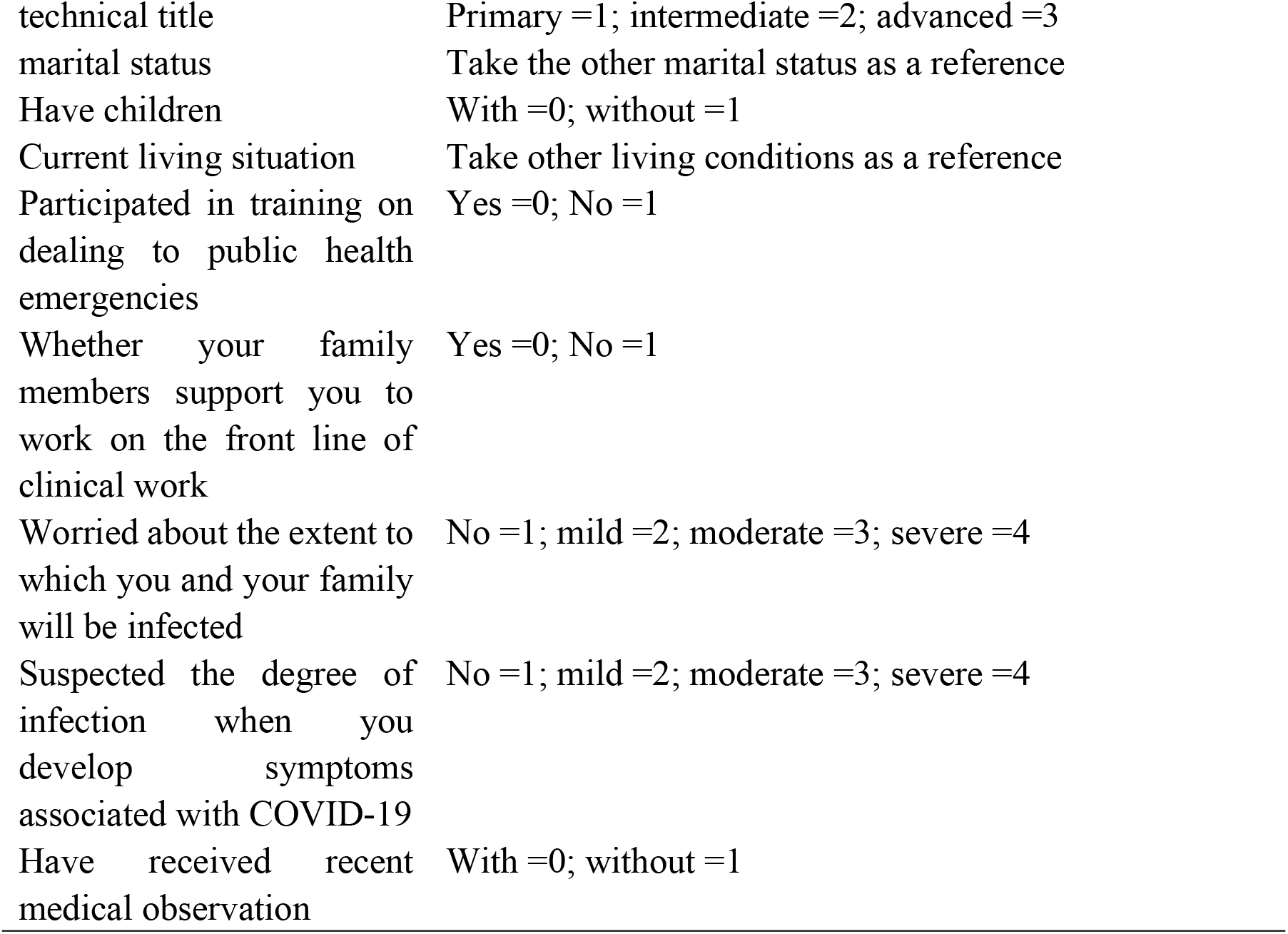
Values of the factors influencing the mental health status of clinical medical staff.

**Table 4.** Results of multiple linear regression analysis of the factors influencing the mental health status of clinical medical staff.

| **variable** | **B** | **Beta** | **t price** | **P price** |
| --- | --- | --- | --- | --- |
| constant | 43.729 |  | 0.064 | 0.949 |
| The extent to which you are infected when you develop symptoms related to COVID-19 | 6.296 | 0.402 | 5.636 | 0.000 |
| Worried about the extent to which you and your family will be infected | 6.804 | 0.323 | 4.573 | 0.000 |
| Have received recent medical observation | 15.457 | -0.146 | -2.334 | 0.021 |
| highest education | 9.458 | 0.133 | 2.099 | 0.038 |

pour: $R^2=0.441$ , $F=28.556$ , $P=0.000$

## Discussion

### Comparison of SCL-90 factor scores and national norm

COVID-19 is a serious infectious disease. Due to its strong infectivity, several people are afraid of it, and even talk about the “crown” color change. This effect reflects on the clinical medical staff. The results of this study showed that the average scores of SCL-90 overall and its somatization, OCD, depressive, anxiety, rival, terror, and psychotic factors were all significantly higher than the normal scores (all P <0.05). This study specifically highlighted that, due to the direct contact with COVID-19 patients, the clinical medical staff represents the population group at the highest risk of COVID-19 infection. At the same time, being those with the deepest understanding of the dangers of COVID-19, they were at increased risk of suffering from anxiety, depression, and fear when they abruptly faced these sudden public health events. The medical staff experienced strong social stress, so their symptoms’ scores on the rating scales were higher than normal, representing a need that prompted the attention of psychologists and team leaders. The clinical medical personnel will be able to better overcome the pandemic only following amelioration of their working conditions and an improved psychological quality for self-regulation and self-protection. The conclusions of this study are partially consistent with those from the assessment of the mental health status of the Severe Acute Respiratory Syndrome (SARS) frontline medical staff performed in 2003 [18].

### Analysis of the factors influencing the mental health status of clinical medical personnel

The results of this study showed that the influencing factors affecting the mental health status were, from high to low: the degree of suspicion of being infected when the symptoms of COVID-19 appeared, the degree of concern for one’s self and one’s family members, a recent medical check-up, and the education level (P <0.05). The specific reasons are analyzed as follows. When presenting COVID-19 related symptoms, the health professionals’ suspicion regarding the extent of the infection and their concern about their own and their families’ infection, the impact of a recent medical check-up is easily understandable.

The main source of the spread of COVID-19 infection are the symptomatic and asymptomatic patients with novel coronavirus infection [19]. When treating and nursing patients, health professionals are in close contact with patients, and their risk of infection is high [20]. Therefore, medical staff is a high-risk group, facing huge psychological pressure. With the emergence of symptoms and the corresponding worry to develop the disease, the mental health status of the health professionals will rapidly deteriorate, greatly affecting their physical and mental health. This, in turn, will translate into a less efficacious treatment of the patients with coronavirus infection that are under their care; therefore, psychological intervention targeted at health professional’s symptoms (related to the context of the COVID-19 pandemic) is warranted and systematic medical/psychological observation of the clinical medical staff is needed in order to relieve their psychological pressure and to promote their recovery as soon as possible.

Regarding the educational level variable in our study, the higher the SCL-90 score, the more the psychological problems of the medical staff. The reason for this trend may be related to the fact that higher-education professionals bear a heavier workload, in terms of professional medical work and technical requirements; competition is also fierce in their field. Highly educated medical professionals are often highly valued by their working unit and by society, they are at the core of hospital clinical, teaching and scientific research tasks. Especially during the pandemic outbreak, they are the pillar of their department, they must participate in the outbreak model, so the physical and mental pressure that they endure is substantially greater than that borne by a medical personnel with less responsibilities and lower-level positions. This is largely consistent with previous studies of mental health status at the time of similar events [19].

### Limitations and further research

The generalizability of this study’s results is limited by its cross-sectional design and by having analyzed convenient samples from front-line medical workers from a single province. Based on this study, future research may use a longitudinal approach with a wider sample to measure the psychological state of front-line medical professionals across multiple dimensions. This approach is expected to identify the interaction between demographics and psychological states in a more comprehensive manner.

## Conclusion

In the face of such a catastrophic emergency as the one represented by the COVID-19 pandemic, the emergence of some psychopathological symptoms in the front-line medical staff is a form of stress response, which is also influenced by various subjective and objective factors. These symptoms can also be seen as an explanatory, emotional, and defensive response processes within the individual, including the body’s physiological response to physical needs or harm. While working in such unique environments, front-line healthcare workers can experience disorders with respect to their work, life, emotions, and overall state. Owing to the requirements for isolation and disinfection, medical workers are required to wear several layers of protective clothing, which adds to their physical labor, consumes a large amount of energy, and leads to a severe lack of oxygen, resulting in physical and psychological symptoms. When confronted with a disaster, people in good mental health would take the initiative to adopt countermeasures, such as speaking out, shifting attention, compensating, relaxing, and turning to humor, self-comfort, and reason. The present results demonstrated that front-line medical workers experience serious psychological problems in their efforts to control the NCP pandemic. The psychological testing showed that people experience a process of adaptation to catastrophic emergencies, from initial unacceptance, shock, and fear, to routine, acceptance, and calmness until co-existence is reached, which is a process with its own order. Facing such sudden disasters as the NCP outbreak, both medical professionals and patients experience psychological clinical symptoms. For front-line medical staff, while ensuring the completion of their duties, keeping good mental health is crucial. Given the special circumstances, it is urgently worth discussing how to strengthen the mental health monitoring of front-line medical workers and establish an active, systematic, and scientific system for their psychological protection.

### Relevance to clinical practice

Our results found that front-line medical staff have psychological problems when dealing with the outbreak of COVID-19 and are presented with considerable psychological stress in the face of the growing number of confirmed cases and the current absence of a specific curative treatment. Consequently, guiding policies and psychological interventions are crucial to maintaining their psychological well-being. Different measures may be implemented to solve this problem. For example, organized rotation and shifts would allow for breaks from working in high-risk areas and facilitate the arrangement of family and social time. Expressed appreciation and support on the part of medical organizations would help prevent burnout and build a resilient team to conserve strength for working in a stressful environment during the outbreak of an emerging infectious disease. Educational programs for medical workers to fulfill their desire for related knowledge and mental health programs to improve their mental well-being are essential and may also translate into a more efficacious management of the infectious disease outbreak.

## Data Availability

Data are available from the Ningbo college of health sciences Ethics Committee for researchers who meet the criteria for access to confidential data.

## Acknowledgments

The authors would like to thank the front-line health workers from different level hospitals in the Zhejiang Province who participated in the study.

## Availability of data and materials

The datasets generated and analyzed during the current study are not publicly available due to ethical restrictions and patient confidentiality but are conditionally available from the corresponding author on request. Aggregated data are provided in the paper tables.

## Authors’ contributions

The authors were responsible for the paper as follows. LYL and NS: conception, design, analysis, and data interpretation, drafting the manuscript, revising the manuscript, and approval of its final version; NS: acquisition of data, project administration, manuscript revisions, and approval its final version; SPZ and CYF: formal analysis, manuscript revision, and final version approval; HYL and SY: conception, manuscript revision, and final version approval; JMX: conception, design, funding acquisition, project administration, manuscript revision, and final version approval. All the authors have read and approved the final manuscript.

## Ethics approval and consent to participate

Permission to conduct the study and to obtain access for the purpose of gathering the data were obtained from by the Ethics Committee of Ningbo College of Health Sciences. Written, informed consent was obtained from all the participants, or from their legal guardians, or caregivers prior to enrolment in the study.

